# Striatal functional connectivity in psychosis relapse: A comparison between antipsychotic adherent and non-adherent patients at the time of relapse

**DOI:** 10.1101/2020.07.07.20148452

**Authors:** Jose M Rubio, Todd Lencz, Anita Barber, Franchesica Bassaw, Gabriela Ventura, Nicole Germano, Anil K Malhotra, John M Kane

**Affiliations:** The Zucker Hillside Hospital, Department of Psychiatry, Northwell Health, Glen Oaks, NY, USA; Zucker School of Medicine at Hofstra/Northwell, Department of Psychiatry and Molecular Medicine, Hempstead, NY, USA; The Feinstein Institute for Medical Research, Center for Psychiatric Neuroscience, Manhasset, NY, USA

## Abstract

Most individuals with psychotic disorders relapse over their course of illness. Relapse pathophysiology is generally not well captured in studies that do not account for antipsychotic non-adherence, which is common and often unnoticed in schizophrenia. This study was explicitly designed to understand relapse in patients with guaranteed antipsychotic delivery. We compared individuals with psychosis breakthrough on antipsychotic maintenance medication (BAMM, n=23), for whom antipsychotic adherence prior to relapse was confirmed by using long acting injectable antipsychotics, and individuals who at the time of relapse were antipsychotic free (APF, n=27), as they had declared treatment non-adherence. Resting state functional MRI was acquired to conduct a region of interest (ROI) analyses. We generated functional connectivity maps to calculate striatal connectivity index (SCI) values, a prognostic biomarker of treatment response in first episode schizophrenia. Group differences in SCI values (BAMM vs APF) were compared in a linear regression model. We hypothesized that individuals in the BAMM group would have greater aberrant striatal function, thus lower SCI values, than in individuals in the APF group. Furthermore, we conducted exploratory group comparisons at the ROI level. As predicted, the BAMM group had significantly lower SCI values (ß=0.95, standard error=0.378, p=0.013). Group comparisons at the ROI level indicate differences in functional connectivity of dorsal striatum, and greater decoupling in striato-cerebellar connections among the BAMM group. A prognostic biomarker of treatment response in first episode psychosis showed differences by antipsychotic exposure upon relapse, suggesting that relapse during continued antipsychotic treatment may be characterized by aberrant striatal function.

---

Most individuals with schizophrenia-spectrum disorders will experience psychosis relapse several times throughout the course of their illness^1^. Relapse is associated with societal and personal burden, is detrimental to recovery, and may represent a danger to self or others^2,3^. Therefore, it is critical to identify the mechanisms involved in psychosis relapse to optimize relapse-prevention strategies and to improve the overall prognosis of psychotic disorders.

The largest contributor to relapse risk is lack of adherence with antipsychotic maintenance treatment^4,5^. Compared with placebo, antipsychotic drugs are highly efficacious in relapse-prevention with a number needed to treat of three^6^. Unfortunately, it is often difficult to disentangle whether relapse occurs due to medication non-adherence, which occurs frequently, or in the context of continued medication delivery. Research on individuals treated with long acting injectable antipsychotics (LAI), for whom continuous antipsychotic exposure is confirmed, overcomes this major confounder^9^. Using this approach, we have previously demonstrated that breakthrough psychosis is relatively common, with an incidence of almost 23 events per 100 participant-years of continuous antipsychotic treatment^10^. This indicates that for a sizeable proportion of patients whose symptoms are stabilized on antipsychotic drugs, these drugs may nevertheless fail to prevent some subsequent exacerbations.

Although research on the mechanisms of psychosis relapse during antipsychotic maintenance treatment is limited, there has been substantial progress in understanding the neural substrate of response to antipsychotic drugs. Measuring resting state functional connectivity (RSFC) of the striatum in individuals with schizophrenia spectrum disorders, several studies converge in finding that striatal RSFC abnormalities prior to treatment onset are associated with treatment response^11–16^. For instance, we previously developed the striatal connectivity index (SCI), a prognostic biomarker derived from the RSFC values from 91 striatal functional connections predictive of treatment response. Individuals with a first psychotic episode who responded to 12 weeks of treatment with risperidone or aripiprazole had lower SCI values than non-responders or healthy controls, a finding which was replicated in an independent cohort^15^. Furthermore, studies on the changes of striatal RSFC over the course of antipsychotic treatment have found a correlation between longitudinal changes in striatal RSFC and symptom improvement^13,16^. These, and additional data derived with other neuroimaging modalities^17–19^, support the theory that greater striatal dysconnectivity (not necessarily disconnectivity per se) before treatment onset predicts treatment response by virtue of being targeted and “stabilized” by antipsychotic drugs in individuals who respond to treatment, whereas individuals with non-response could have other functional deficits not targeted by current antipsychotic drugs^18^.

The closest relevant data for how these findings could translate to relapse prevention derives from animal models. In a series of experiments of acute and chronic antipsychotic treatment in rats, Samaha et al demonstrated how haloperidol and olanzapine over time lost their ability to suppress amphetamine-induced locomotion and conditioned avoidance response, and how this is related to compensatory changes in the dopamine (DA) system associated with chronic DA 2 receptor (D2R) blockade. In particular, what they observed is that this loss of “efficacy” of antipsychotic drugs over time was not related to changes in the availability of striatal DA, but rather an increment in the density and affinity of postsynaptic D2R over time^20^.

Taken together, these data suggest that psychosis relapse despite continuous antipsychotic treatment could at least partly result from dynamic changes in the dopaminergic system with chronic antipsychotic exposure. These changes would disturb the stabilization of striatal functional connectivity previously achieved by antipsychotic drugs, ultimately leading to the recurrence of striatal dysconnectivity and worsening of psychotic symptoms.

Therefore, in this study, we aimed to test aspects of this theory by measuring the striatal RSFC in patients who relapsed despite guaranteed medication delivery with LAIs, versus a group of patients who relapsed while non-adherent to treatment. Our primary hypothesis was that the failure of continuous antipsychotic treatment to prevent psychosis relapse would be associated with greater aberrant striatal function, thus lower SCI values, than in individuals who at the time of relapse were not treated with antidopaminergic drugs. Furthermore, we conducted additional group comparisons of RSFC in various striatal sub-regions, to identify the specific connections contributing to group differences in SCI values.

## METHODS

The study design consisted of a cross-sectional comparison of striatal RSFC measured with functional MRI (fMRI), between individuals with multiepisode psychosis spectrum disorder who were experiencing psychosis “breakthrough on antipsychotic maintenance medication” (BAMM group), and individuals with the same diagnosis who were “antipsychotic free” at the time of relapse (APF group) (Figure 1).

**Figure 1.**
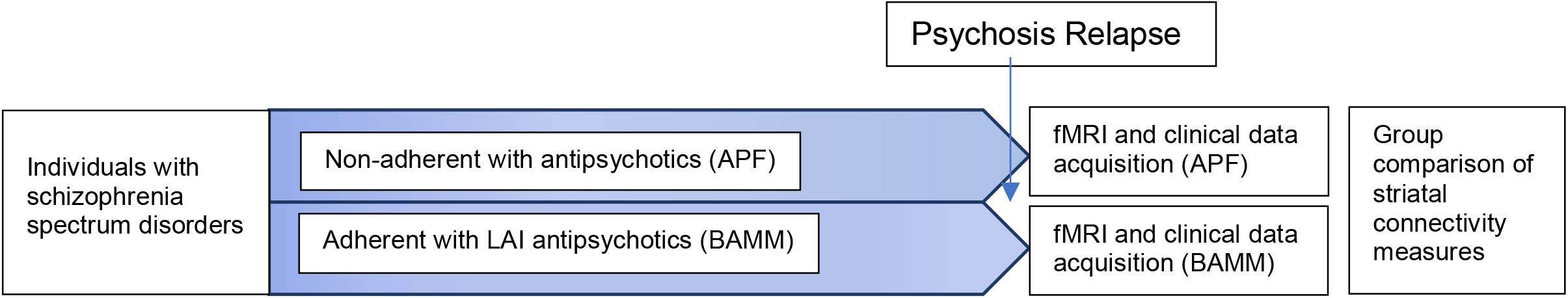
Study design.

### Participants

Patients aged 18-65 receiving treatment at The Zucker Hillside Hospital – Northwell Health with a chart diagnosis of a psychotic disorder or bipolar disorder with psychotic features with at least one previous episode, who presented for treatment of symptom exacerbation were assessed with the Structural Clinical Interview for DSM-IV (SCID)^21^ and the Brief Psychiatric Rating Scale - Anchored (BPRS-A)^22^, and were enrolled in the study if they met the following criteria: 1) diagnosis of schizophrenia, schizoaffective disorder, psychotic disorder not otherwise specified or bipolar I disorder with psychotic features, 2) No psychiatric hospitalization within previous 3 months, 3) Current positive symptoms rated ≥4 (moderate) on one or more of these Brief Psychiatric Rating Scale - Anchored (BPRS-A)^22^ items: conceptual disorganization, grandiosity, hallucinatory behavior, unusual thought content. Additional criteria were used to divide these individuals into the groups that were compared in the study. Individuals participating in the APF group needed to: 1) Be non-adherent with antipsychotic drugs prior to the worsening of their symptoms according to the clinical assessment conducted upon arrival to the hospital, 2) Have a medication log which reflected that no antipsychotic was administered between hospital admission and time of the scan, and 3) Confirmation by participant to study personnel antipsychotic of non-adherence between symptom worsening and time of scan. Alternatively, individuals participating in the BAMM group needed to have medical record documentation that they were on active treatment with a LAI antipsychotic, and that this treatment had been continuous, for at least 3 months prior to the time of the scan.

Antipsychotic exposure at the time of the scan was confirmed by testing the plasma level of the LAI antipsychotic being prescribed for the BAMM group, and for the APF group the most likely antipsychotic to last be prescribed (either the last antipsychotic that the patient had access to at home or haloperidol, which is the most frequently used in the emergency room for agitation). Plasma samples were sent to the Analytical Psychopharmacology Laboratory of the Nathan Kline Institute in Orangeburg, NY, where plasma levels of olanzapine, risperidone, paliperidone, aripiprazole, haloperidol or fluphenazine were measured using validated liquid chromatographic methods^8^. To determine whether antipsychotic plasma levels were therapeutic we followed parameters of the Arbeitsgemeinschaft für Neuropsychopharmakologie und Pharmakopsychiatrie (AGNP) expert group consensus guidelines for therapeutic drug monitoring (TDM), allowing for a maximum of 10% below the lower threshold, since these have indicative purposes only and there is not a correlation between plasma level and efficacy. The cutoffs were 20-60ng/dl for paliperidone, 1-10ng/dl for haloperidol, 0.8-10 ng/dl for fluphenazine, 20-80ng/dl for olanzapine and 100-350ng/dl for aripiprazole^23,24^.

In addition to diagnostic and psychotic symptom severity assessments, we conducted clinical assessments of negative, depressive^25^, and manic symptoms^26^, as well as of known risk factors for relapse such as stressful life events^27^, and resiliency^28^, and urine toxicology status at the time of the scan.

All patients signed informed consent, and all procedures were approved by the Institutional Review Board (IRB) of the Feinstein Institutes for Medical Research – Northwell Health.

### Resting State fMRI Image Acquisition and Preprocessing

Resting state fMRI (rs-fMRI) scans were collected on a 3T Siemens Prisma scanner utilizing a multi-band accelerated echo-planar imaging (EPI) sequence described in detail in the Human Connectome Project ^29^. For each study participant, we acquired a T1-weighted scan (TR=2400 msec, TE=2.22 msec, voxel size=0.8 mm3, scan length=6 min, 38 s) and two 7-minute 17-second rsMRI runs, one each with AP and PA phase encoding directions. The first 13 volumes were discarded acquisitions. Resting scans contained 594 whole-brain volumes, each with 72 contiguous axial/oblique slices in the AC-PC orientation (TR=720ms, TE=33.1ms, matrix = 104×90, FOV = 208mm, voxel = 2×2×2mm, multi-band acceleration factor=8). During the scans, participants were instructed to stay awake with their eyes closed and to think of nothing in particular.

The neuroimaging preprocessing methods first corrected the 3D T1 images for scanner-dependent gradient field non-linearities using a gradient unwarp tool^30^. Standard structural preprocessing was then done according to the HCP preprocessing pipelines which included gradient distortion correction, brain extraction, cross-modal registration of T2 weighted (T2w) images to T1w, bias field correction based on square root (T1w*T2w) and non-linear registration to MNI space^31^. The functional preprocessing methods used were gradient distortion correction, motion correction, and EPI image distortion correction based on spin-echo EPI field maps (FSL toolbox “topup”), and spatial registration to T1w image and MNI space^31^. An initial high pass filter of 2000 Hz was applied to remove any slow drift trends before nuisance regression was performed using FMRIB’s ICA-based X-noiseifier (FIX)^32–34^. Functional images then underwent 5-mm full-width-at-half-maximum spatial smoothing and temporal bandpass-filtering (0.1-0.01 Hz). Frame-wise Displacement (FD) was calculated for each scan time point and any scans with FD exceeding 0.5 mm were removed from further analysis^35^. All participants included in the final study sample had at least 10 minutes of usable resting-state scan data after scrubbing.

### Statistical and RSFC Analyses

In order to calculate the SCI, we first measured the RSFC of subregions of the striatum, using a seed-based approach. Regions of interest (ROI) within the striatum were defined as in the original Di Martino et al. study^36^, which has been subsequently used in studies of antipsychotic treatment response^15,16,37^. Bilateral 3.5mm spherical ROIs were located in dorsal caudate (DC) (x = ±13, y = 15, and z = 9), ventral striatum superior (VSs) (x = ±10, y = 15, and z = 0), ventral striatum inferior (VSi) (x = ±9, y = 9, and z = −8), dorsal rostral putamen (DRP) (x = ±25, y = 8, and z = 6), dorsal caudal putamen (DCP) (x = ±28, y = 1, and z = 3), and the ventral rostral putamen (VRP) (x = ±20, y = 12, and z = −3). After defining the 12 ROIs, we extracted their mean time course of the resting state blood oxygen level dependent (BOLD) signal for each subject. Whole-brain voxel wise correlation maps for each ROI were created with the extracted waveform as a reference, and the resulting correlation maps were z-transformed. Connectivity maps resulting from the different phase encoding directions (i.e., AP and PA) were averaged to obtain one connectivity map per seed and scan. Whether global signal regression (GSR) should be regressed-out of the time-series for each voxel remains as controversial topic, since although this approach may introduce artifactual anti-correlations^38^, not all anti-correlations found in GSR analyses are artifactual, and in fact this approach may show better signal in system specific correlations and show better correspondence with the anatomy by removing non-neural contributions to the BOLD signal.^39,40^ Therefore, we ran the analyses with and without GSR, interpreting that given our interest in system-specific correlations (i.e., striatal connectivity), GSR is probably most appropriate, and that concordance between GSR and No GSR results would reflect most consistency.

Once we had generated connectivity maps or each phase encoding direction with and without GSR, we proceeded to calculate the SCI for each of them, following a similar approach as in previous research^15,41^. Briefly, we extracted the 91 striatal functional connections that were used to calculate the SCI in the original Sarpal et al study^15^, and applied to those the same weights as in the original study, to compute a SCI values per scan session in each phase encoding direction (i.e., AP vs PA), which were later averaged into a single SCI value per scan (i.e, study participant), generating a SCI value using GSR and another using No GSR. Next, these SCI values were entered into a linear regression model adjusting for sex and age, in which group status (i.e., APF vs BAMM) was entered as covariate of interest. Differences were deemed statistically significant at p<0.05. SCI value calculations and analyses were conducted with the R Studio version 1.2.5019^42^. Data and code to generate these results are available on https://github.com/lorente01/psychosisrelapseRSFC.

Finally, we conducted exploratory analyses to identify the connections with greatest differences in RSFC between the two groups. For this, we used SPM12 (https://www.fil.ion.ucl.ac.uk/spm/software/spm12/) called from Matlab 2015b. Connectivity maps for each group (APF and BAMM) were visually inspected, and we found a good separation of networks, consistent with the results of the Di Martino et al study^36^. For each one of the ROIs we set up a generalized linear model, using group (i.e., BAMM vs APF) as contrast of interest, with sex, age, FD-DVARS correlation, and scan duration after scrubbing as regressors. For these analyses, we used a voxel-level threshold of p<0.01, with cluster level threshold of p<0.05 corrected for false discovery rate (FDR)^43^ by the standard function provided by the SPM12 package.

## RESULTS

### Sample characteristics

50 participants were included in the analyses, 23 in the BAMM group and 27 in the APF group. The mean age was 34.97 years (Standard Deviation [SD]=12.77), and half of the sample was female, with no significant differences between groups (p=0.07 and p>0.9 respectively). At the time of relapse, the mean BPRS was 42.59 (SD=7.23), and the psychotic sub-score of the BPRS was 14.08 (SD=3.17). There were no significant differences between groups in psychotic (p=0.7), negative (p=0.3), manic (p=0.09) or depressive symptoms (p>0.9). There was no significant difference between groups either in current stressful life events (p=0.07, resilience (p=0.2) or positive urine toxicology screen (p=0.2). The mean duration of resting state fMRI acquired post-scrubbing was 13.35 minutes (SD=1.04), with no difference between groups (p=0.2). In the BAMM group, the LAIs prescribed upon relapse were aripiprazole (n=7; 30%), paliperidone (n=9, 39%), fluphenazine decanoate (n=1;4.3%), haloperidol decanoate (n=6, 26%) (Table 1).

**Table 1.** Participant characteristics.

|  | Total Sample<br>(n=50)<br>N (%) | By Group |  | p Value |
| --- | --- | --- | --- | --- |
|  |  | BAMM (n=23)<br>N (%) | APF (n=27)<br>N (%) |  |
| Gender |  |  |  | >0. |
| Female | 25 (50%) | 12 (52%) | 13 (48%) |  |
| Male | 25 (50%) | 11 (48%) | 14 (52%) |  |
|  | **Mean (SD)** | **Mean (SD)** | **Mean (SD)** |  |
| Age (years) | 34.97 (12.77) | 39.92 (15.22) | 30.75 (8.44) | 0.07 |
| Laterality Quotient | 77.31 (39.06) | 83.13 (14.93) | 74.10 (47.56) | 0. |
| Body Mass Index | 27.84 (7.75) | 29.67 (7.21) | 26.40 (8.00) | 0. |
| Brief Psychopathology Rating Scale Psychotic Subscore | 14.08 (3.17) | 13.91 (3.42) | 14.23 (3.00) | 0. |
| Brief Psychopathology Rating Scale Score | 42.59 (7.23) | 42.67 (7.79) | 42.52 (6.89) | >0. |
| Negative Symptom Assessment Score | 3.34 (1.12) | 3.53 (1.07) | 3.20 (1.15) | 0. |
| Calgary Depressive Symptom Scale Score | 2.31 (10.37) | 1.47 (12.31) | 2.88 (9.05) | >0. |
| Young Mania Rating Scale Score | 16.18 (5.75) | 14.80 (6.44) | 17.28 (5.00) | 0.08 |
| Connor Davidson Resiliency Scale Score | 64.69 (25.70) | 57.50 (26.22) | 69.48 (24.82) | 0. |
| Holmes Rahe Stressful Life Events Scale Score | 185.28 (153.30) | 127.71 (102.68) | 227.83 (171.87) | 0.06 |
| Mean resting state minutes acquired | 13.35 (1.04) | 13.05 (1.30) | 13.60 (0.68) | 0 |
|  | **N (%)** | **N (%)** | **N (%)** |  |
| Positive urine toxicology screen | 11 (31%) | 2 (15%) | 9 (41%) | 0. |
| LAI Antipsychotic prescribed and plasma level |  |  |  |  |
| Aripiprazole | 7 (14%) | 7 (14%) | 0 |  |
| Aripiprazole plasma level (Mean, SD; ref 100-350 ng/dl) |  | 137.6 (97.85) | ND |  |
| Paliperidone | 9 (18%) | 9 (18%) | 0 |  |
| Paliperidone plasma level (Mean, SD; ref 20-60 ng/dl) |  | 36.63 (8.14) | ND |  |
| Fluphenazine | 1 (2.0%) | 1 (2.0%) | 0 |  |
| Fluphenazine plasma level (Mean, SD; ref 0.8-10 ng/dl) |  | 2.4 (NA) | ND |  |
| Haloperidol | 6 (12%) | 6 (12%) | 0 |  |
| Haloperidol plasma level (Mean, SD; ref 1-10 ng/dl) |  | 7.5 (3.87) | ND |  |

### Differences in the SCI between relapse during ongoing antipsychotic treatment and during antipsychotic non-adherence

We found that the SCI values in the BAMM group were significantly lower than for individuals in the APF group, both in GSR and No GSR analyses. Specifically, the difference between groups for GSR calculated SCI were ß=0.95, standard error=0.378, p=0.013, and the for SCI calculated with No GSR were ß=1.317, standard error=0.643, p=0.046 (Figure 2). The correlation coefficient between GSR derived SCI and No GSR derived SCI was r=0.74.

**Figure 2.**
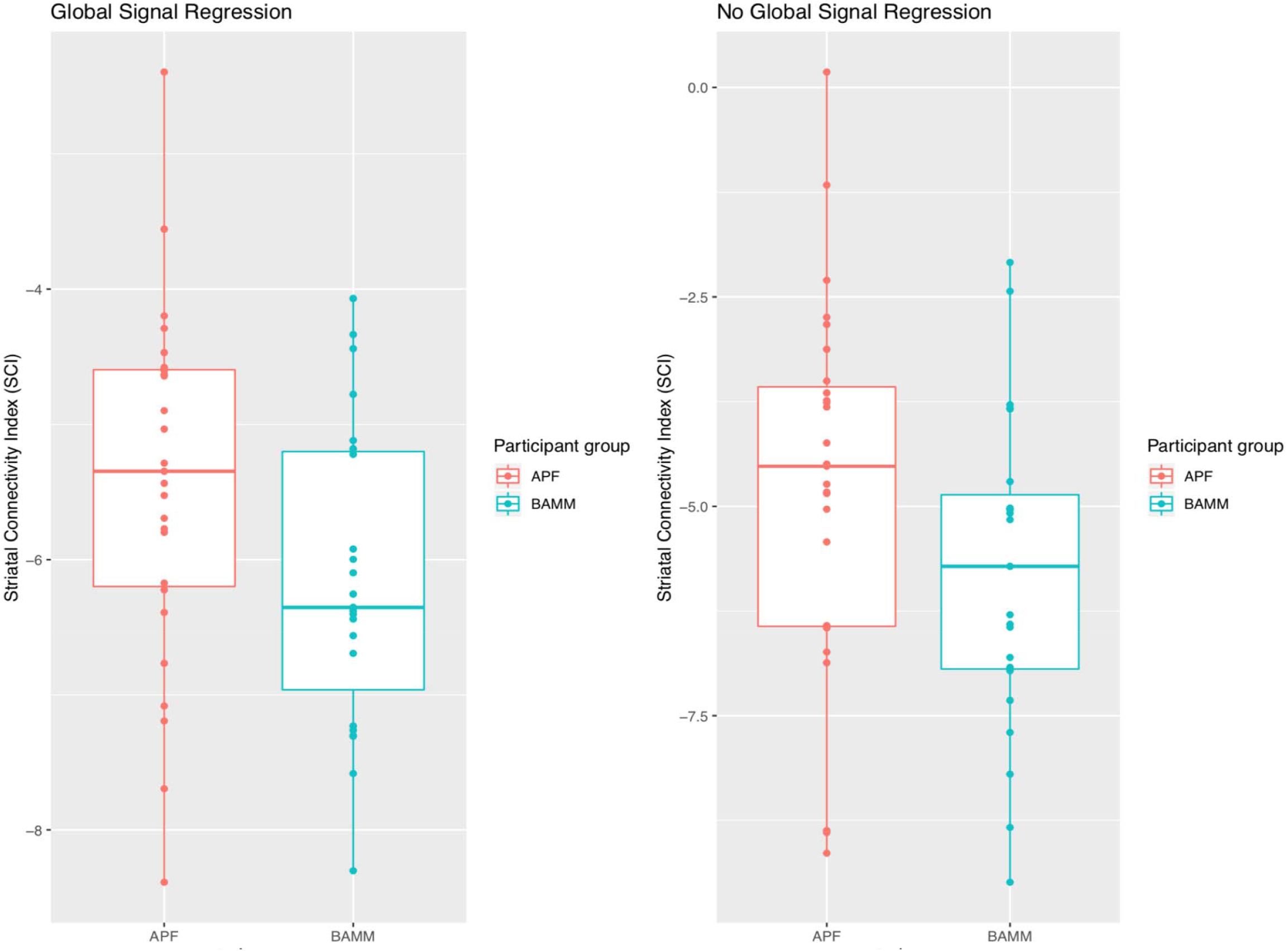
Striatal Connectivity Index (SCI) value upon psychosis relapse by antipsychotic exposure status. Note: Comparison of SCI value between APF and BAMM groups ß=-0.95025, p Value=0.0130 in analyses with GSR and ß =-1.3167, p Value=0.0464 in analyses with No GSR adjusted for sex and age.

### Differences in striatal ROI functional connectivity between relapse during ongoing antipsychotic treatment and during antipsychotic non-adherence

In our exploratory analyses, we identified 27 functional connections for which there were significant group differences for results with GSR, and 8 for results without GSR. Most of the ROI for which there were group differences were in the dorsal striatum (DC and DCP). In both the GSR (FDR corrected p Value=0.001, T value=4.47) and No GSR analyses (FDR corrected p Value=0.035, T value=3.81), the left dorsal caudate was hyperconnected with the middle temporal gyrus in the BAMM compared with the APF group. Also, consistently between GSR and No GSR analyses, there was lower functional connectivity in striato-cerebellar functional connections in the BAMM group than in the APF group (DCR and DCL in GSR analyses, DCPL in No GSR analyses) (Table 2 and Figure 3).

**Table 2.** Significant Differences in Striatal Functional Connectivity by Antipsychotic Treatment Status at Relapse.

| Global Signal Regression |  |  |  |  |  |  |  |  |  | No Global Signal Regression |  |  |  |  |  |  |  |  |  |
| --- | --- | --- | --- | --- | --- | --- | --- | --- | --- | --- | --- | --- | --- | --- | --- | --- | --- | --- | --- |
| Seed | Contrast Direction | p Value* | K Size † | T Value | Z Value | MNI Coordinates |  |  | Region | Seed | Contrast Direction | p Value* | K Size | T Value | Z Value | MNI Coordinates |  |  | Region |
|  |  |  |  |  |  | x | y | z |  |  |  |  |  |  |  | x | y | z |  |
| DCL | BAMM>APF | 0.001 | 219 | 4.47 | 4.03 | 58 | -62 | 12 | Middle Temporal Gyrus | DCL | BAMM>APF | 0.035 | 152 | 3.81 | 3.52 | 54 | -62 | 12 | Middle Temporal Gyrus |
|  | APF>BAMM | <0.001 | 418 | 5.36 | 4.66 | -32 | -76 | -28 | Cerebellum posterior lobe | DCPL | APF>BAMM | <0.001 | 365 | 5.28 | 4.61 | 0 | -94 | 8 | Visual Cortex |
|  |  | 0.03 | 109 | 4.99 | 4.4 | -34 | 14 | 50 | Supplementary motor area |  |  | 0.011 | 171 | 4.96 | 4.39 | 54 | -60 | -34 | Cerebellum |
|  |  | 0.039 | 100 | 4.75 | 4.24 | -12 | -70 | 52 | Superior parietal lobule |  |  | 0.031 | 135 | 3.63 | 3.37 | -36 | -68 | -28 | Cerebellum |
|  |  | <0.001 | 247 | 4.59 | 4.12 | 40 | -56 | -48 | Cerebellum posterior lobe | VRPR | APF>BAMM | <0.001 | 429 | 4.58 | 4.11 | -48 | 40 | 20 | Middle Frontal Gyrus |
|  |  | 0.002 | 179 | 4.52 | 4.07 | -18 | -82 | -30 | Cerebellum posterior lobe |  |  | <0.001 | 303 | 4.58 | 4.11 | -44 | -44 | 48 | Inferior Parietal Lobule |
|  |  | <0.001 | 228 | 4.3 | 3.9 | 6 | -82 | -24 | Cerebellum posterior lobe |  |  | 0.019 | 158 | 4.47 | 4.03 | 36 | -60 | 44 | Angular Gyrus |
|  |  | 0.041 | 96 | 3.96 | 3.64 | 0 | -40 | 30 | Posterior cingulate |  |  | 0.01 | 184 | 4.29 | 3.89 | -50 | -50 | -14 | Middle Temporal Gyrus |
| DCR | BAMM>APF | <0.001 | 695 | 4.88 | 4.33 | 8 | -86 | 36 | Cuneus |  |  |  |  |  |  |  |  |  |  |
|  |  | <0.001 | 227 | 4.49 | 4.04 | 48 | -20 | 56 | Postcentral gyrus |  |  |  |  |  |  |  |  |  |  |
|  |  | 0.007 | 147 | 4.16 | 3.79 | 50 | -70 | 8 | Middle Temporal Gyrus |  |  |  |  |  |  |  |  |  |  |
|  | APF>BAMM | <0.001 | 518 | 5.4 | 4.69 | -38 | -78 | -42 | Cerebellum posterior lobe |  |  |  |  |  |  |  |  |  |  |
|  |  | <0.001 | 503 | 5.29 | 4.61 | 36 | -64 | -46 | Cerebellum posterior lobe |  |  |  |  |  |  |  |  |  |  |
|  |  | <0.001 | 202 | 5.1 | 4.49 | -14 | -82 | -22 | Cerebellum posterior lobe |  |  |  |  |  |  |  |  |  |  |
|  |  | <0.001 | 534 | 4.87 | 4.32 | -40 | -58 | 42 | Inferior parietal lobule |  |  |  |  |  |  |  |  |  |  |
|  |  | 0.003 | 146 | 4.86 | 4.31 | -52 | -48 | -42 | Cerebellum posterior lobe |  |  |  |  |  |  |  |  |  |  |
|  |  | <0.001 | 641 | 4.84 | 4.3 | -46 | 32 | 28 | Middle Frontal gyrus |  |  |  |  |  |  |  |  |  |  |
|  |  | 0.001 | 168 | 4.54 | 4.08 | 58 | -46 | -4 | Middle Temporal Gyrus |  |  |  |  |  |  |  |  |  |  |
|  |  | <0.001 | 258 | 4.51 | 4.06 | 36 | -58 | 44 | Inferior parietal lobule |  |  |  |  |  |  |  |  |  |  |
|  |  | <0.001 | 299 | 4.5 | 4.05 | -26 | 6 | 62 | Middle Frontal gyrus |  |  |  |  |  |  |  |  |  |  |
| VRPL | BAMM>APF | 0.007 | 161 | 4.66 | 4.17 | -10 | -52 | 48 | Precuneus |  |  |  |  |  |  |  |  |  |  |
|  |  | 0.001 | 245 | 4.42 | 3.99 | 20 | -50 | 2 | Parahippocampal Gyrus |  |  |  |  |  |  |  |  |  |  |
|  |  | 0.047 | 101 | 3.86 | 3.55 | -14 | -48 | 0 | Visual associative cortex |  |  |  |  |  |  |  |  |  |  |
|  |  | 0.047 | 103 | 3.65 | 3.39 | 52 | -46 | 14 | Angular gyrus |  |  |  |  |  |  |  |  |  |  |
| VRPR | BAMM>APF | 0.003 | 205 | 4.82 | 4.29 | -2 | -56 | 46 | Posterior cingulate |  |  |  |  |  |  |  |  |  |  |
| VSIL | APF>BAMM | 0.013 | 155 | 4.09 | 3.74 | 40 | -48 | 59 | Postcentral gyrus |  |  |  |  |  |  |  |  |  |  |
| VSSL | BAMM>APF | 0.03 | 137 | 4.04 | 3.7 | 54 | -8 | 4 | Auditory cortex |  |  |  |  |  |  |  |  |  |  |

Note: \* p Value corrected for false discovery rate ; † Size of 2x2x2mm voxels in cluster. Legend: MNI="Montréal Neurological Institute"; BAMM="Breakthrough on Antipsychotic Maintenance Medication group"; APF="Antipsychotic Free group"; DCL="Left Dorsal Caudate"; DCR="Right Dorsal Caudate"; VRPL="Left Ventrorostral Putamen"; VRPR="Right Ventrorostral Putamen"; VSIL="Left Ventroinferior Striatum"; VSSL="Left Ventrosuperior Striatum"; DCPL="Left Dorsocaudal Putamen".

**Figure 3.**
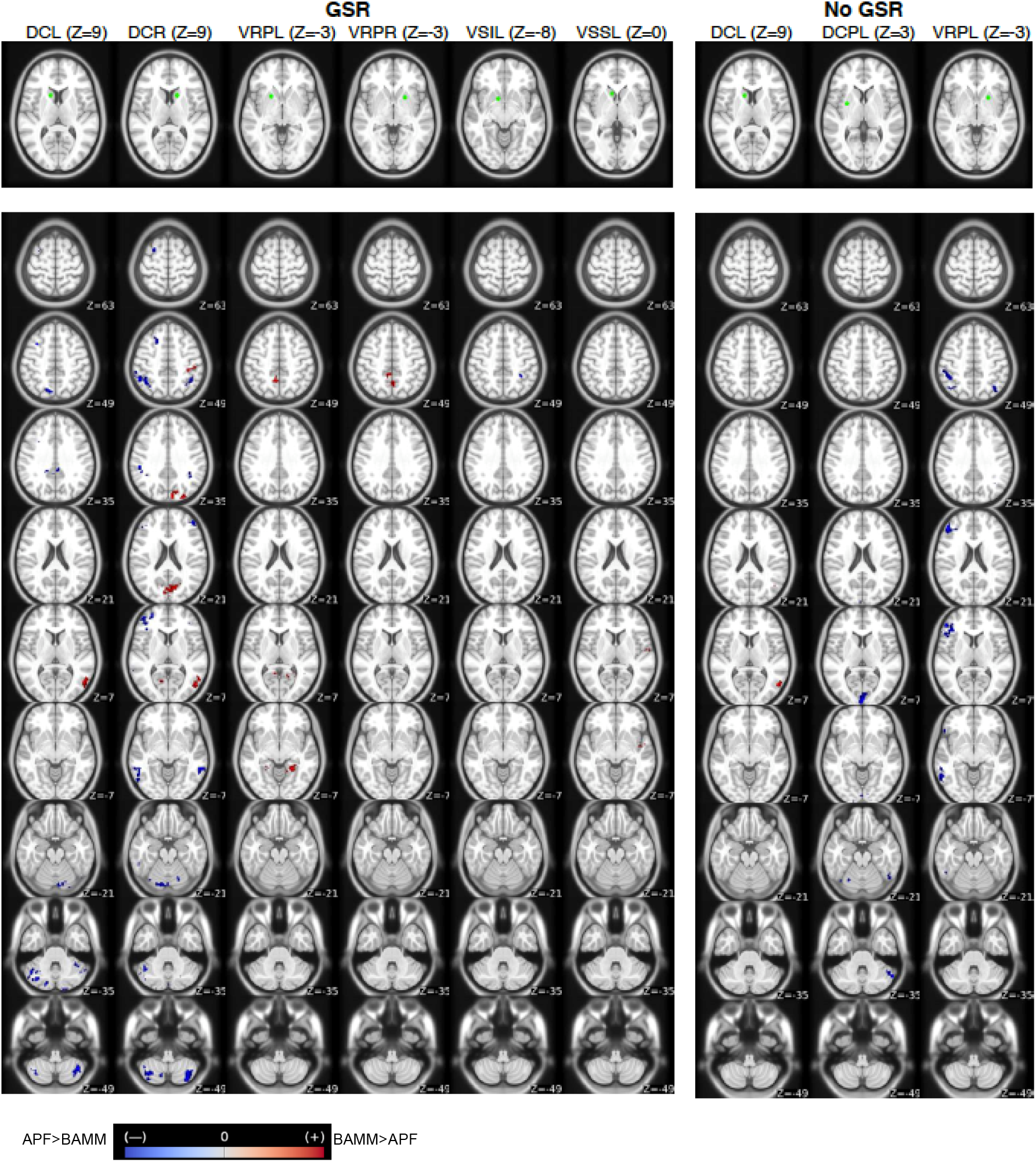
Differences in RSFC between groups by striatal region of interest. **Legend:** DCL=Left dorsal caudate; DCR=Right dorsal caudate; VRPL=Left ventrorostral putamen; VRPR=Right ventrorostral putamen; VSIL Left ventral striatum inferior; VSSL= Left ventral striatum superior. Green colors reflect the location of each region of interest. Warmer colors reflect increased RSFC in BAMM than in APF, whereas colder colors represent increased RSFC in APF than BAMM. Z refers to the axial pla in the MNI coordinate system.

## DISCUSSION

To our knowledge, this is the first functional neuroimaging study of psychosis relapse explicitly designed to remove the confounder of antipsychotic treatment non-adherence. We expand on prior work on the application of the SCI as a prognostic biomarker of antipsychotic response in first episode psychosis^15^, and the effect of cannabis use on treatment response,^41^ to psychosis relapse. As predicted by our hypothesis, the SCI values were significantly lower for individuals whose symptom worsening occurred despite ongoing antipsychotic treatment (i.e., BAMM) than for those who had discontinued antipsychotic drugs prior to relapse (i.e., APF). This finding aligns with our theory that the recurrence of striatal dysfunction during ongoing antipsychotic treatment would have a causal role in psychosis relapse, and as such, SCI values that normalized with treatment would now return to the same values as when symptomatic, despite continued antipsychotic exposure.

Our findings are informative about the behavior of the SCI and striatal functional connectivity as illness progresses after the first treatment with antipsychotic drugs. In our exploratory analyses, we found group differences at the level of some of the 91 functional connections that make up the SCI. For instance, individuals in the BAMM group had greater RSFC between VRPR and posterior cingulate than individuals in the APF group. RSFC in this connection is predictive of treatment response in first episode patients,^15^ hence driving the SCI in a negative direction (lower SCI values predict treatment response). This exemplifies how compared to individuals who relapsed without antipsychotic treatment, the BAMM group had overall greater RSFC among the 91 functional connections which positively predict treatment response in first episode patients, but lower for those functional connections negatively predictive of response in first episode patients. This shows that rather than a general pattern of decoupling between the striatum and cortical regions, dysconnectivity in the BAMM group was featured by both hyper and hypoconnectivity in those meaningful 91 connections, compared with individuals who relapsed off antipsychotics.

Group differences in SCI values were statistically significant when calculated with and without global signal regression, with a moderate to high correlation between both methods. Yet, although the general direction of the results was similar between GSR and No GSR for the region of interest analyses, the were a number of distinct connections for which there were group differences between these methods. This is expected, as system-wide measurements, such as the SCI, are less sensitive to removing the global signal than system-specific measurements, such as region of interest analyses, for which GSR would be preferred^40^. Still, we found some overlap between both approaches for some specific connections (e.g., greater RSFC in BAMM than APF for functional connectivity between DCL and middle temporal gyrus), in a preponderance of group differences in dorsal striatal regions, and in a consistent lower striato-cerebellar functional connectivity for individuals in the BAMM group.

Dorsal striatal loops process primarily motor information^44^, yet a growing body of literature shows that dopaminergic dysfunction in this striatal division, for which the main dopaminergic input is the nigrostriatal pathway^45^, is core to the dopaminergic dysfunction of schizophrenia, rather than in the mesolimbic pathway as previously thought^17,45–47^. The accumulating literature on the dorsal (motor) striatum as the locus of dopaminergic dysfunction in psychosis in general,^47^ and in our case in psychosis relapse during antipsychotic treatment in particular, aligns with recent relevant clinical observations. In an individual participant data meta-analysis of psychosis relapse-prevention clinical trials with LAI antipsychotics, the strongest predictor of relapse was tardive dyskinesia (TD), with a 239% increment in risk^10^. The pathophysiology of TD involves reorganization of monoaminergic (mostly DA and 5HT)^20,48–50^ function in motor domains of the striatum resulting from chronic antipsychotic exposure^51^, and in fact the only approved treatment for this condition are VMAT2 inhibitors^52^, which essentially decrease the presynaptic monoaminergic release in the striatum^53^. The strong predictive effect of TD on relapse during antipsychotic treatment, thought to be mediated by dorsostriatal dysfunction, and our cross-sectional finding of the greatest dysconnectivity between dorsal striatum and associative cortical and cerebellar areas at the time of relapse during antipsychotic treatment, support the hypothesis that changes resulting from chronic dopaminergic exposure, particularly in nigrostriatal pathways, may be involved in the pathophysiology of psychosis relapse during antipsychotic treatment. Investigation of dysfunction in this pathway with methods such as neuromelanin sensitive MRI^45,54^, is warranted to test this hypothesis

Similar to cortico-striatal loops, the cortico-cerebellar loops are topographically organized by the type of information that they process, and once thought to work in parallel with cortico-striatal circuits, striatal and cerebellar systems do have anatomical and functional direct connections and are in functional balance with each other.^55^ Striato-cerebellar functional connections are decoupled in schizophrenia compared with healthy controls^56^, and in our study such decoupling was greater when relapse occurred during ongoing antipsychotic treatment (i.e., BAMM). Cerebellar functional connectivity abnormalities have been involved in cognitive^57^ and negative symptoms^58^ in chronically treated patients. Our study design is not sufficient to discriminate whether the finding of striato-cerebellar decoupling is driven by pathophysiological differences between relapse on vs off antipsychotic, by effects of antipsychotic drugs on connectivity independent of their clinical effects, or by group differences in cognition. Subsequent longitudinal study designs should be able to disentangle these factors.

Our findings add complexity to the dichotomy of striatal vs extra-striatal dysfunction that has been proposed as a model for the pathophysiology of treatment response and resistance in psychosis^18,19^. According to this theory, whereas striatal dopaminergic dysfunction would be a critical element in the pathophysiology of psychosis in “treatment responsive” individuals, extra-striatal mechanisms would mediate the psychotic symptoms in “treatment non-responsive”. The results of this study suggest that striatal mechanisms may be involved in the inability of antipsychotic drugs to prevent subsequent relapses, and that while extra-striatal mechanisms have been identified in treatment resistance,^59,60^ dynamic factors related to the compensatory response to chronic antipsychotic exposure in the dopaminergic system may constitute an additional mechanism. It is plausible that such mechanistic heterogeneity explains why the literature on the mechanisms of treatment response has converged showing striatal dysfunction as a common pathophysiological element^14,61–66^, but it has proven far more elusive to isolate mechanisms of antipsychotic treatment resistance^67,68^

Several limitations should be considered in the interpretation of these data. First, due to its cross-sectional design, this study cannot confirm that the group differences that we found were driven only by changes over time in striatal functioning, hence longitudinal replication of these findings is necessary. Second, given the nature of the comparison (i.e., relapse on vs off antipsychotics) we could not completely disentangle what differences were driven by distinct pathophysiological mechanisms between relapse on and off antipsychotics versus the effects of antipsychotic drugs on striatal RSFC independent from their clinical effects. Third, our overall hypothesis assumes that individuals who relapsed during ongoing antipsychotic treatment previously had shown clinical improvement with that same treatment. In prior research we demonstrated that this applies to at least half of individuals with relapse during ongoing antipsychotic treatment^10^, and in this study entry criteria included 12 weeks of clinical stability prior to relapse; however, future longitudinal research should prospectively demonstrate treatment response prior to relapse.

As a first foray into the functional connectivity features of psychosis relapse, our results point towards future directions in this area of research. First, longitudinal fMRI study designs, and comparison of individuals with relapse during ongoing antipsychotic treatment with symptom stability during continuous antipsychotic treatment seem the next steps to confirm some of our findings. Second, the group differences in the SCI and the finding of striato-cerebellar decoupling in BAMM, may correspond with system-wide functional dysconnectivity in BAMM beyond the striatum^56^. Future research should characterize the extent to which the observed differences here expand beyond striatal functioning into resting state networks. Finally, following up on preclinical data,^20^ future clinical and translational research should study the dynamic response of monoaminergic systems to chronic antipsychotic exposure, and its implications in striatal functional connectivity, as these would be potential treatment targets for psychosis relapse prevention.

In conclusion, in the first neuroimaging study of psychosis relapse explicitly designed to overcome the confounder of non-adherence with antipsychotic drugs, we found that a prognostic biomarker for treatment response in first episode schizophrenia was significantly different between individuals who relapsed on antipsychotics vs off antipsychotics. This finding aligns with the theory that recurrent striatal dysconnectivity, driven by adaptive changes to chronic antipsychotic exposure, has an effect in the recurrence of psychotic symptoms despite ongoing antipsychotic treatment. Future research should continue testing the various elements of this theory.

## Data Availability

Data and code to generate these results are available on https://github.com/lorente01/psychosisrelapseRSFC.

## Acknowledgments

We want to acknowledge the study participants and their families, as well as Dr Suckow and Cooper for the antipsychotic plasma quantification.

## Author contributions

Study design: JR, TL, AKM, JMK

Data collection: JR, GV, FB, NG

Analyses: JR, AB

Manuscript: All authors

## References

1. Kahn, R. S. et al. Schizophrenia. Nat. Rev. Dis. Primer 1, 15067 (2015).

2. Andreasen, N. C., Liu, D., Ziebell, S., Vora, A. & Ho, B.-C. Relapse duration, treatment intensity, and brain tissue loss in schizophrenia: a prospective longitudinal MRI study. Am. J. Psychiatry 170, 609– 615 (2013).

3. Pennington, M. & McCrone, P. The Cost of Relapse in Schizophrenia. PharmacoEconomics 35, 921– 936 (2017).

4. Robinson, D. et al. Predictors of relapse following response from a first episode of schizophrenia or schizoaffective disorder. Arch. Gen. Psychiatry 56, 241–247 (1999).

5. Alvarez-Jimenez, M. et al. Risk factors for relapse following treatment for first episode psychosis: a systematic review and meta-analysis of longitudinal studies. chizophr. Res. 139, 116–128 (2012).

6. Leucht, S. et al. Antipsychotic drugs versus placebo for relapse prevention in schizophrenia: a systematic review and meta-analysis. Lancet Lond. Engl. 379, 2063–2071 (2012).

7. Kane, J. M., Kishimoto, T. & Correll, C. U. Non-adherence to medication in patients with psychotic disorders: epidemiology, contributing factors and management strategies. World Psychiatry 12, 216– 226 (2013).

8. Lopez, L. V. et al. Accuracy of Clinician Assessments of Medication Status in the Emergency Setting: A Comparison of Clinician Assessment of Antipsychotic Usage and Plasma Level Determination. J. Clin. Psychopharmacol. 37, 310–314 (2017).

9. Rubio, J. M. & Kane, J. M. Psychosis breakthrough on antipsychotic maintenance medication (BAMM): what can we learn? NPJ Schizophr. 3, 36 (2017).

10. Rubio, J. Breakthrough antipsychotic maintenance medication: An individual participant data metaanalysis of individuals adherent with long acting injectables. http://yoda.yale.edu/table-3-data-requests-approved.

11. Li, A. et al. A neuroimaging biomarker for striatal dysfunction in schizophrenia. Nat. Med. 26, 558– 565 (2020).

12. Li, H. et al. Enhanced baseline activity in the left ventromedial putamen predicts individual treatment response in drug-naive, first-episode schizophrenia: Results from two independent study samples. EBioMedicine 46, 248–255 (2019).

13. Han, S. et al. Distinct striatum pathways connected to salience network predict symptoms improvement and resilient functioning in schizophrenia following risperidone monotherapy. Schizophr. Res. 215, 89–96 (2020).

14. Doucet, G. E., Moser, D. A., Luber, M. J., Leibu, E. & Frangou, S. Baseline brain structural and functional predictors of clinical outcome in the early course of schizophrenia. Mol. Psychiatry 25, 863–872 (2020).

15. Sarpal, D. K. et al. Baseline Striatal Functional Connectivity as a Predictor of Response to Antipsychotic Drug Treatment. Am. J. Psychiatry 173, 69–77 (2016).

16. Sarpal, D. K. et al. Antipsychotic Treatment and Functional Connectivity of the Striatum in First-Episode Schizophrenia. JAMA Psychiatry 72, 5 (2015).

17. Howes, O. D. et al. The nature of dopamine dysfunction in schizophrenia and what this means for treatment. Arch. Gen. Psychiatry 69, 776–786 (2012).

18. Howes, O. D. & Kapur, S. A neurobiological hypothesis for the classification of schizophrenia: type A (hyperdopaminergic) and type B (normodopaminergic). Br. J. Psychiatry J. Ment. Sci. 205, 1–3 (2014).

19. Howes, O. D., McCutcheon, R., Owen, M. J. & Murray, R. M. The Role of Genes, Stress, and Dopamine in the Development of Schizophrenia. Biol. Psychiatry 81, 9–20 (2017).

20. Samaha, A.-N., Seeman, P., Stewart, J., Rajabi, H. & Kapur, S. ‘Breakthrough’ dopamine supersensitivity during ongoing antipsychotic treatment leads to treatment failure over time. J. Neurosci. Off. J. Soc. Neurosci. 27, 2979–2986 (2007).

21. First MB, Williams JBW, Karg RS, Spitzer RL. Structured Clinical Interview for DSM-5—Research Version (SCID-5 for DSM-5, Research Version; SCID-5-RV). (American Psychiatric Association; US, 2015).

22. Woerner, M. G., Mannuzza, S. & Kane, J. M. Anchoring the BPRS: an aid to improved reliability. Psychopharmacol. Bull. 24, 112–117 (1988).

23. Noel, C. A review of a recently published guidelines’ ‘strong recommendation’ for therapeutic drug monitoring of olanzapine, haloperidol, perphenazine, and fluphenazine. Ment. Health Clin. 9, 287– 293 (2019).

24. Hiemke, C. et al. Consensus Guidelines for Therapeutic Drug Monitoring in Neuropsychopharmacology: Update 2017. Pharmacopsychiatry 51, 9–62 (2018).

25. Addington, D., Addington, J. & Maticka-Tyndale, E. Assessing depression in schizophrenia: the Calgary Depression Scale. Br. J. Psychiatry. Suppl. 39–44 (1993).

26. Young, R. C., Biggs, J. T., Ziegler, V. E. & Meyer, D. A. A rating scale for mania: reliability, validity and sensitivity. Br. J. Psychiatry J. Ment. Sci. 133, 429–435 (1978).

27. Noone, P. A. The Holmes-Rahe Stress Inventory. Occup. Med. Oxf. Engl. 67, 581–582 (2017).

28. Connor, K. M. & Davidson, J. R. T. Development of a new resilience scale: the Connor-Davidson Resilience Scale (CD-RISC). Depress. Anxiety 18, 76–82 (2003).

29. Van Essen, D. C. et al. The Human Connectome Project: a data acquisition perspective. NeuroImage 62, 2222–2231 (2012).

30. Jovicich, J. et al. Reliability in multi-site structural MRI studies: effects of gradient non-linearity correction on phantom and human data. NeuroImage 30, 436–443 (2006).

31. Glasser, M. F. et al. The minimal preprocessing pipelines for the Human Connectome Project. NeuroImage 80, 105–124 (2013).

32. Smith, S. M. et al. Resting-state fMRI in the Human Connectome Project. NeuroImage 80, 144–168 (2013).

33. Griffanti, L. et al. ICA-based artefact removal and accelerated fMRI acquisition for improved resting state network imaging. NeuroImage 95, 232–247 (2014).

34. Salimi-Khorshidi, G. et al. Automatic denoising of functional MRI data: combining independent component analysis and hierarchical fusion of classifiers. NeuroImage 90, 449–468 (2014).

35. Power, J. D., Schlaggar, B. L. & Petersen, S. E. Recent progress and outstanding issues in motion correction in resting state fMRI. NeuroImage 105, 536–551 (2015).

36. Di Martino, A. et al. Functional connectivity of human striatum: a resting state FMRI study. Cereb. Cortex N. Y. N 1991 18, 2735–2747 (2008).

37. Sarpal, D. K. et al. Relationship between Duration of Untreated Psychosis and Intrinsic Corticostriatal Connectivity in Patients with Early Phase Schizophrenia. Neuropsychopharmacol. Off. Publ. Am. Coll. Neuropsychopharmacol. 42, 2214–2221 (2017).

38. Murphy, K., Birn, R. M., Handwerker, D. A., Jones, T. B. & Bandettini, P. A. The impact of global signal regression on resting state correlations: are anti-correlated networks introduced? NeuroImage 44, 893–905 (2009).

39. Fox, M. D., Zhang, D., Snyder, A. Z. & Raichle, M. E. The global signal and observed anticorrelated resting state brain networks. J. Neurophysiol. 101, 3270–3283 (2009).

40. Murphy, K. & Fox, M. D. Towards a consensus regarding global signal regression for resting state functional connectivity MRI. NeuroImage 154, 169–173 (2017).

41. Blair Thies, M. et al. Interaction of Cannabis Use Disorder and Striatal Connectivity in Antipsychotic Treatment Response. Schizophr. Bull. Open 1, sgaa014 (2020).

42. RStudio Team. RStudio Team (2019). RStudio: Integrated Development for R. (2019).

43. Chumbley, J., Worsley, K., Flandin, G. & Friston, K. Topological FDR for neuroimaging. NeuroImage 49, 3057–3064 (2010).

44. Alexander, G. E., DeLong, M. R. & Strick, P. L. Parallel organization of functionally segregated circuits linking basal ganglia and cortex. Annu. Rev. Neurosci. 9, 357–381 (1986).

45. Cassidy, C. M. et al. Neuromelanin-sensitive MRI as a noninvasive proxy measure of dopamine function in the human brain. Proc. Natl. Acad. Sci. 116, 5108–5117 (2019).

46. McCutcheon, R. A., Abi-Dargham, A. & Howes, O. D. Schizophrenia, Dopamine and the Striatum: From Biology to Symptoms. Trends Neurosci. 42, 205–220 (2019).

47. McCutcheon, R., Beck, K., Jauhar, S. & Howes, O. D. Defining the Locus of Dopaminergic Dysfunction in Schizophrenia: A Meta-analysis and Test of the Mesolimbic Hypothesis. Schizophr. Bull. 44, 1301–1311 (2018).

48. Charron, A., Hage, C. E., Servonnet, A. & Samaha, A.-N. 5-HT2 receptors modulate the expression of antipsychotic-induced dopamine supersensitivity. Eur. Neuropsychopharmacol. 25, 2381–2393 (2015).

49. Lévesque, C. et al. Deficient striatal adaptation in aminergic and glutamatergic neurotransmission is associated with tardive dyskinesia in non-human primates exposed to antipsychotic drugs. Neuroscience 361, 43–57 (2017).

50. Blin, J. et al. Striatal dopamine D2 receptors in tardive dyskinesia: PET study. J. Neurol. Neurosurg. Psychiatry 52, 1248–1252 (1989).

51. Teo, J. T., Edwards, M. J. & Bhatia, K. Tardive dyskinesia is caused by maladaptive synaptic plasticity: a hypothesis. Mov. Disord. Off. J. Mov. Disord. Soc. 27, 1205–1215 (2012).

52. Solmi, M., Pigato, G., Kane, J. M. & Correll, C. U. Treatment of tardive dyskinesia with VMAT-2 inhibitors: a systematic review and meta-analysis of randomized controlled trials. Drug Des. Devel. Ther. 12, 1215–1238 (2018).

53. DeJesus, O. T., Shelton, S. E., Roberts, A. D., Nickles, R. J. & Holden, J. E. Effect of tetrabenazine on the striatal uptake of exogenous L-DOPA in vivo: a PET study in young and aged rhesus monkeys. Synap. N. Y. N 44, 246–251 (2002).

54. Wengler, K., He, X., Abi-Dargham, A. & Horga, G. Reproducibility assessment of neuromelanin-sensitive magnetic resonance imaging protocols for region-of-interest and voxelwise analyses. NeuroImage 208, 116457 (2020).

55. Bostan, A. C. & Strick, P. L. The basal ganglia and the cerebellum: nodes in an integrated network. at. Rev. Neurosci. 19, 338–350 (2018).

56. Ji, J. L. et al. Schizophrenia Exhibits Bi-directional Brain-Wide Alterations in Cortico-Striato-Cerebellar Circuits. Cereb. Cortex 29, 4463–4487 (2019).

57. Andreasen, N. C. et al. Schizophrenia and cognitive dysmetria: a positron-emission tomography study of dysfunctional prefrontal-thalamic-cerebellar circuitry. Proc. Natl. Acad. Sci. 93, 9985–9990 (1996).

58. Brady, R. O. et al. Cerebellar-Prefrontal Network Connectivity and Negative Symptoms in Schizophrenia. Am. J. Psychiatry 176, 512–520 (2019).

59. Demjaha, A. et al. Antipsychotic treatment resistance in schizophrenia associated with elevated glutamate levels but normal dopamine function. Biol. Psychiatry 75, e11–13 (2014).

60. Iwata, Y. et al. Glutamatergic Neurometabolite Levels in Patients With Ultra-Treatment-Resistant Schizophrenia: A Cross-Sectional 3T Proton Magnetic Resonance Spectroscopy Study. Biol. Psychiatry 85, 596–605 (2019).

61. Dandash, O., Pantelis, C. & Fornito, A. Dopamine, fronto-striato-thalamic circuits and risk for psychosis. Schizophr. Res. 180, 48–57 (2017).

62. Eisenberg, D. P. et al. Presynaptic Dopamine Synthesis Capacity in Schizophrenia and Striatal Blood Flow Change During Antipsychotic Treatment and Medication-Free Conditions. Neuropsychopharmacol. Off. Publ. Am. Coll. Neuropsychopharmacol. 42, 2232–2241 (2017).

63. Fornito, A. et al. Functional dysconnectivity of corticostriatal circuitry as a risk phenotype for psychosis. JAMA Psychiatry 70, 1143–1151 (2013).

64. Horga, G. & Abi-Dargham, A. The striatum and dopamine: a crossroad of risk for schizophrenia. JAMA Psychiatry 71, 489–491 (2014).

65. Jauhar, S. et al. Determinants of treatment response in first-episode psychosis: an 18F-DOPA PET study. Mol. Psychiatry (2018) doi:10.1038/s41380-018-0042-4.

66. Jauhar, S. et al. The Effects of Antipsychotic Treatment on Presynaptic Dopamine Synthesis Capacity in First-Episode Psychosis: A Positron Emission Tomography Study. Biol. Psychiatry 85, 79–87 (2019).

67. Nakajima, S. et al. Neuroimaging findings in treatment-resistant schizophrenia: A systematic review: Lack of neuroimaging correlates of treatment-resistant schizophrenia. Schizophr. Res. 164, 164–175 (2015).

68. Mouchlianitis, E., McCutcheon, R. & Howes, O. D. Brain-imaging studies of treatment-resistant schizophrenia: a systematic review. Lancet Psychiatry 3, 451–463 (2016).

